# Will Proton Pump Inhibitors Lead to a Higher Risk of COVID-19 Infection and Progression to Severe Disease? A Meta-analysis

**DOI:** 10.1101/2020.12.25.20248860

**Authors:** Cunye Yan, Yue Chen, Chenyu Sun, Mubashir Ayaz Ahmed, Chandur Bhan, Ce Cheng, Lei Hu, Zhichun Guo, Hongru Yang, Chenyu Cao, Ziwei Ji, Yue Yan, Yijing Zuo, Yiceng Sun, Yao Li, Qin Zhou

## Abstract

**Background:** Previous researches on the association between proton pump inhibitors (PPIs) use and the treatment and prevention of COVID-19 have generated inconsistent findings. Therefore, this Meta-analysis was conducted to clarify the outcome in patients who take PPIs.

**Methods:** We carried out a systematic search to identify potential studies until November 2020. Heterogeneity was assessed using the I-squared statistic. Odds ratios (ORs) with its 95% confidence intervals (CIs) were calculated by fixed-effects or random-effects models according to the heterogeneity. Sensitivity analyses and tests for publication bias were also performed.

**Results:** Eight articles with more than 268,683 subjects were included. PPI use was not associated with increased or decreased risk of COVID-19 infection (OR:3.16, 95%CI = 0.74-13.43, *P*=0.12) or mortality risk of COVID-19 patients (OR=1.91, 95% CI=0.86-4.24, *P*=0.11). While it can add risk of severe disease (OR=1.54, 95% CI=1.20-1.99, *P*<0.001;) and secondary infection (OR=4.33, 95% CI=2.57-7.29). No publication bias was detected.

**Conclusions:** PPI use is not associated with increased risk infection and may not change the mortality risk of COVID-19, but appeared to be associated with increased risk of progression to severe disease and secondary infection. However, more original studies to further clarify the relationship between PPI and COVID-19 are still urgently needed.

## Introduction

In March 2020, the World Health Organization (WHO) announced that Coronavirus Disease 2019 (COVID-19) would become a pandemic in 2020^[1]^. As of November 15, 2020, more than 53.5 million people have been diagnosed with COVID-19 and 1.3 million people have died of the virus^[2]^, whereas the pandemic is not yet under control. This global pandemic has seriously affected public health and socio-economic development around the world. Therefore, it is of great significance to identify the risk factors of COVID-19 infection, as well as proper treatment modalities. In previous studies, it was found that severe acute respiratory syndrome coronavirus 2 (SARS-CoV-2), the virus causing COVID-19, enters the human body not only through the respiratory system, but also through the gastrointestinal system^[3]^. The virus can rapidly invade and replicate intestinal cells using angiotensin-converting enzyme-2 receptor, which is widely expressed in the GI tract^[4]^. It has been reported that nearly half of COVID-19 patients have viral RNA in their feces^[5]^. These results suggested a strong correlation between gastrointestinal system and SARS-CoV-2 infection. Therefore, it is of great importance to study the relationship between the most commonly used acid suppressor, proton pump inhibitors (PPI), and COVID-19 infection. PPI was postulated to be beneficial in the treatment and prevention of COVID-19^[6]^, while other evidence suggested that PPI could potentially aggravate the condition of patients with COVID-19 and increase the mortality rate^[7]^. As previous studies reached controversial conclusions, we conducted this meta-analysis to further explore the relationship between PPI use and COVID-19, with more detailed subgroup analyses on different outcome index.

## Methods

This meta-analysis was reported in conformity to the Preferred Reporting Items for Systematic Reviews and Meta-Analyses (PRISMA) ^[8]^. And it was registered on International Prospective Register of Systematic Reviews (PROSPERO).

### Literature search strategy

The articles were collected until November 2020 from PubMed, Google scholar, Embase, medRxiv, Web of Science, CNKI, Wanfang and VIP database. The keywords were (“PPI” OR “PPIs” OR “proton pump inhibitors” OR “proton pump inhibitor” OR “omeprazole” OR “lansoprazole” OR “dexlansoprazole” OR “esomeprazole” OR “pantoprazole” OR “rabeprazole” OR “acid suppress” OR “acid suppression”) AND (“COVID-19” OR “Novel coronavirus” OR “coronavirus disease” OR “SARS-CoV-2” OR “Severe acute respiratory syndrome coronavirus 2”). In addition, reference lists of the retrieved articles were reviewed to identify other eligible studies..

### Identification of eligible studies

Two authors (C. Yan and C. Sun) independently carried out the search and disagreements were solved by consensus. The abstract of the retrieved studies was reviewed initially and excluded if deemed irrelevant. The full text of the relevant studies was further reviewed for eligibility. If there were duplicate publications of the same study, the one with the most detailed information and complete data was included.

Studies included in this meta-analysis must meet all of the following criteria: (1) The type of study must be either a randomized controlled trial or an observational study (including cohort and case-control studies). (2) The research was conducted to investigate COVID-19 outcomes, including infection risk, progression to severe cases, death, and risk of secondary infection. (3) PPI use was the exposure for the experimental group and any feasible intervention without PPI use can be implemented on the control group. (4) Relative risks (RRs), hazard ratios (HRs), or odds ratios (ORs) with corresponding 95% confidence intervals (CIs) were able to be extracted or recalculated. (5) The studies were published in English or Chinese. The exclusion criteria were set for this study. (1) The type of articles cannot be accurately determined (2) No valid account of ending data can be derived from the article. (3) Duplicate articles (4) Reviews, case report, animal studies or in vitro studies.

### Data extraction and quality evaluation

Each of the two authors (C. Yan and C. Sun) independently disposed the data from the literature, and if discrepancies arose, a consensus was reached by consulting a third person (Y. Chen), comprehensively comparing the data and through conferral. Information was collected as follows: first author, publication year, country, study design, outcome of the research, NOS scores, the number of cases and controls, odds ratios (ORs) with corresponding 95% confidence intervals (CIs).

Methodological quality of the observational researches was appraised using a validated Newcastle-Ottawa Scale. Three broad subscales including study group selection (0 to 4 points), the groups comparability (0 to 2 points), and the exposures and outcomes elucidation (0 to 3 points). A score of 4-6 was considered moderate, and a score of 7 or more is defined as high quality. Two evaluators (C. Yan and Y. Chen) independently conducted the quality assessment and if there was a divergence of opinion, it would be resolved by mutual communication.

### Statistical analysis and publication bias evaluation

RevMan software (version 5.3; Cochrane Library) and STATA statistical Software (version 14.0; StataCorp, College Station, TX) were utilized for statistical analysis. The associations between PPI use and risk of COVID-19 infection, progression to severe disease, death or risk of secondary infection were analyzed through pooling the ORs and corresponding 95% CIs.

Heterogeneity tests were performed based on Q test and I^2^ statistics. For *I*^*2*^ >50%, w the random effects model was implemented to create forest plots. In contrast, the fixed-effects model was adopted if *I*^*2*^ was <50%. Sensitivity analysis was executed by sequentially excluding the incorporated studies and observing whether the combined results changed significantly. In addition, we conducted a series of subgroup analyses in accordance with the distinct characteristics of the different studies. Publication bias was assessable on a funnel plot qualitatively and Begg’s test and Egger’s test quantitatively. All p-values were two-tailed and *P* < 0.05 was considered statistically significant.

## Results

### Study selection and Study characteristics

**Figure 1** gives the outline of inclusion process, a final electronic search identified 600 records. After eliminating duplicate literature, screening titles and abstracts, we selected 127 potentially relevant articles for further screening, among which, 36 full-text were assessed for eligibility. Eventually, we identified 14 articles published by November 2020, comprising a total of more than 268,683 subjects, including 7 cohort studies and 7 case-control studies. Detailed characteristics of the included studies are showed in **Table 1**. All studies are of a moderate or high quality.

**Table 1.** Main characteristics of studies included in this meta-analysis

| First Author | Year | Country | Study design | Outcomes | PPI use | Non-PPI use | NOS |
| --- | --- | --- | --- | --- | --- | --- | --- |
|  |  |  |  |  | *n* (%) | *n* (%) | score |
| Almario | 2020 | America | Case-control | Morbidity; | 16547 | 36583 | 7 |
| Argenziano | 2020 | America | Case-control | Severity; | 163 | 837 | 8 |
| Cheung | 2020 | China | Cohort | Severity; | 4 | 47 | 9 |
| Corcoles | 2020 | Spain | Cohort | Morbidity; | 99 | 11708 | 8 |
| Freedberg | 2020 | America | Cohort | Severity; | N/A | N/A | 8 |
| Huh | 2020 | South Korea | Case-control | Morbidity; | 660 | 14167 | 8 |
| Jimenez | 2020 | Brazil | Cohort | Severity; | N/A | N/A | 6 |
| Lee | 2020 | Korean | Cohort | Morbidity; Severity; | 20405 | 111911 | 8 |
| Li | 2020 | China | Case-control | Morbidity; Infection; | 95 | 143 | 8 |
| Luxemburger | 2020 | Germany | Case-control | Severity; Mortality; Infection; | 62 | 90 | 7 |
| McKeigue | 2020 | Britain | Case-control | Severity; | 2378 | 34013 | 8 |
| Ramachandran | 2020 | America | Cohort | Severity; Mortality; | 46 | 249 | 7 |
| Tarlow | 2020 | America | Case-control | Morbidity; | 355 | 2535 | 6 |
| Ullah | 2020 | Britain | Cohort | Morbidity; Mortality; | 5908 | 9678 | 8 |

N/A: Not applicable

**Figure 1.**
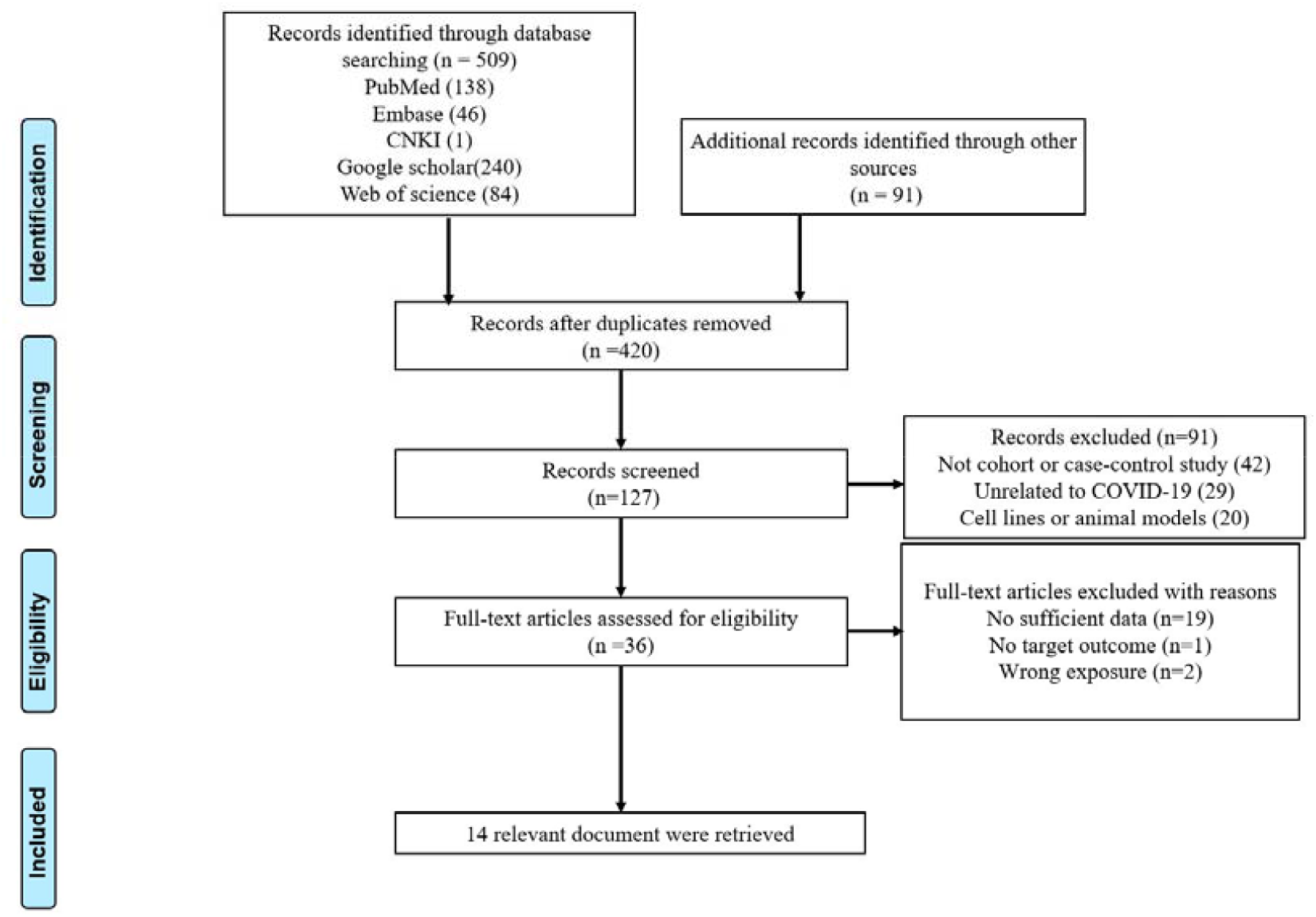
PRISMA Flow Chart

### Meta-analysis

#### Association between PPI use and risk of COVID-19 infection

Analysis of seven studies^[9-15]^ on risk of COVID-19 infection found that the use of PPI was not associated with increased or decreased risk of COVID-19 infection (OR:1.64, 95%CI = 0.54-5.00, *P*=0.390; *I*^*2*^=100%, *P*_*heterogeneity*_ <0.001). (**Figure 2**) In subgroup analysis by study designs, no statistically significant association was observed in cohort (OR:1.19, 95%CI = 0.79-1.79, *P*=0.420; *I*^*2*^=89%, *P*_*heterogeneity*_ =0.001) and case-control studies (OR:2.08, 95%CI = 0.36-12.13, *P*=0.410; *I*^*2*^=100%, *P*_*heterogeneity*_ <0.001). Regarding different geographic locations, no statistically significantassociation was observed in western countries (OR:1.70, 95%CI = 0.33-8.67, *P*=0.520; *I*^*2*^=100%, *P*_*heterogeneity*_ <0.001) and Asian countries (OR:1.30, 95%CI = 0.74-2.29, *P*=0.360; *I*^*2*^=97%, *P*_*heterogeneity*_ <0.001). When it comes to sensitive analysis, studies were omitted one by one. The combined result did not change significantly when any of the articles was excluded. Additionally, no publication bias was detected by Begg’s test (z=0.600, *P*=0.548) and Egger’s test (t=-0.440, *P*=0.677).

**Figure 2.**
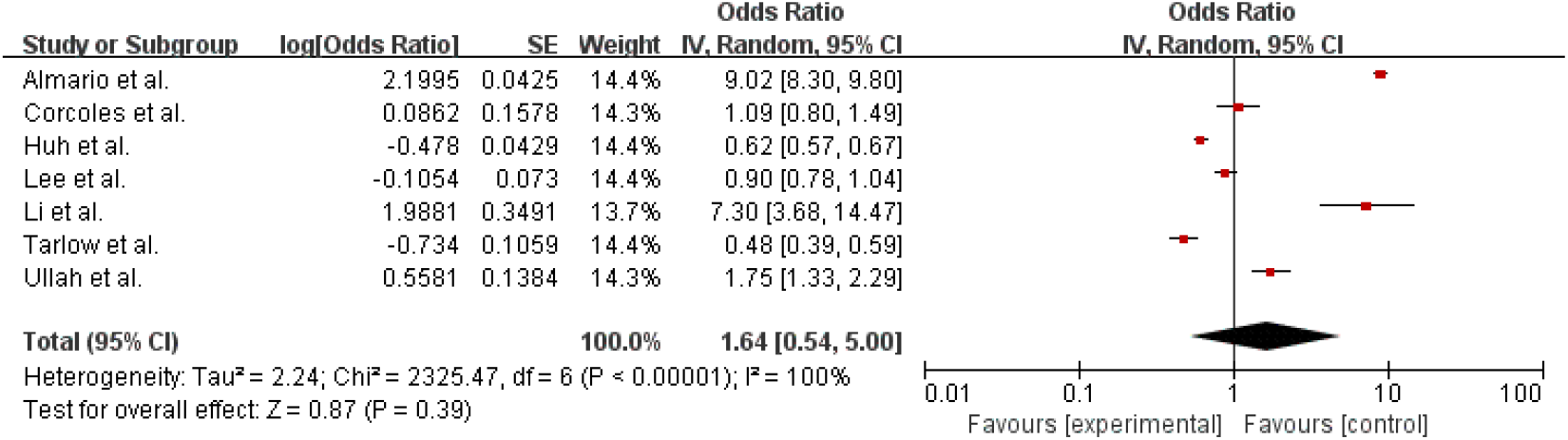
Forest plot: Association between PPI use and risk of COVID-19 infection

#### Association between PPI use and risk of severe disease for COVID-19 patients

The result of nine studies from eight original articles^[7,9, 16-21]^ showed that PPI use was associated with increased risk of severe disease, including admission to ICU, intubation, and death, for COVID-19 patients (OR=1.67, 95% CI=1.37-2.02, *P*<0.001; *I*^*2*^=67%, *P*_*heterogeneity*_=0.002) (**Figure 3**). In subgroup analysis by study designs, a non-significant increased risk was observed in cohort studies(OR:1.39, 95%CI = 0.96-2.01, *P*=0.080; *I*^*2*^=65%, *P*_*heterogeneity*_ =0.060), and a statistically significant increased risk was found for case-control studies (OR:1.86, 95%CI = 1.46-2.37, P=0.410; *I*^*2*^=56%, *P*_*heterogeneity*_ =0.040). Regarding different geographic locations, increased risks were found in both western countries (OR:1.67, 95%CI = 1.33-2.09, *P*<0.001; *I*^*2*^=73%, *P*_*heterogeneity*_=0.001) and Asian countries (OR:1.75, 95%CI = 1.28-2.41, *P*<0.001; *I*^*2*^=0%, *P*_*heterogeneity*_ =0.420). The same method as above was used for sensitivity analysis. The pooled OR has been fluctuating between 1.58 and 1.77 with lower limit of 95% CI constantly remained more than 1 and the *P* value has always been less than 0.05. On top of that, neither Begg’s test (z=0.310, *P*=0.754) nor Egger’s test (t=1.050, *P*=0.327) exhibited evidential publication bias.

**Figure 3.**
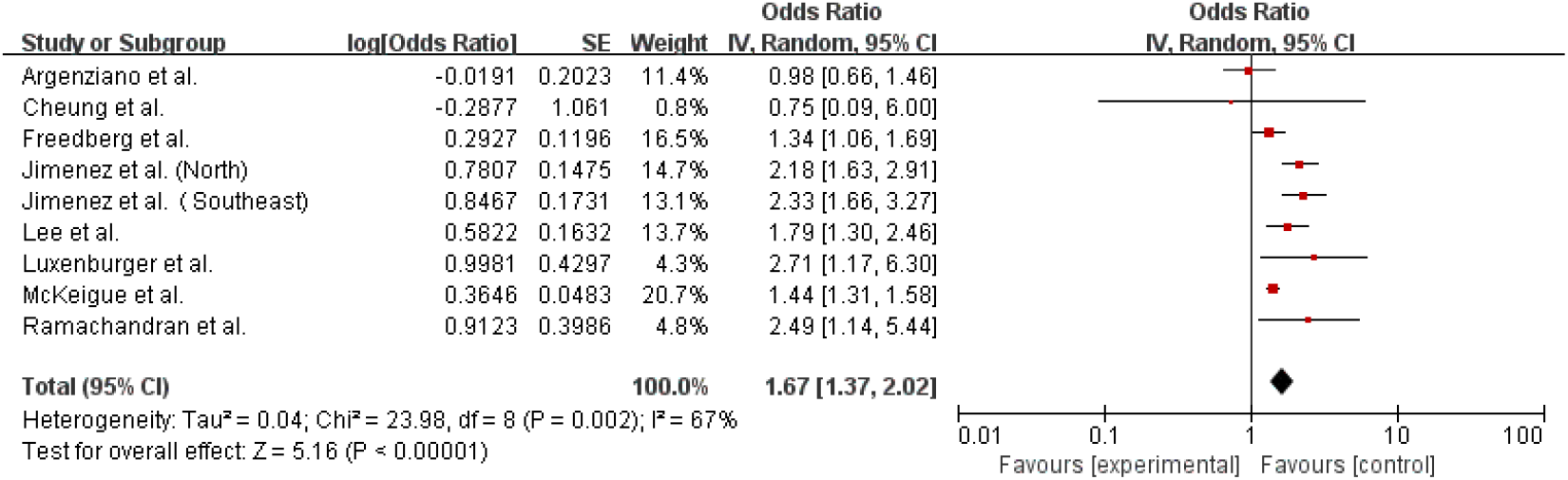
Forest plot: Association between PPI use and risk of severe disease for COVID-19 patients.

#### Association between PPI use and mortality risk for COVID-19 patients

The results indicated no association between PPI use and increased or decreased mortality risk for COVID-19 patients (OR=1.91, 95% CI=0.86-4.24, *P*=0.11; *I*^*2*^=73%, P_heterogeneity_ =0.030) based on three included studies^[7,11,17]^ **(Figure 4)**. Significant change in pooled OR and heterogeneity were found after excluding the article by Ullah et al^[11]^. Furthermore, no publication bias was suspected when three relative studies were emerged. (Begg’s test (z=1.040, *P*=0.296) and Egger’s test (t=1.78, *P*=0.325).

**Figure 4.**
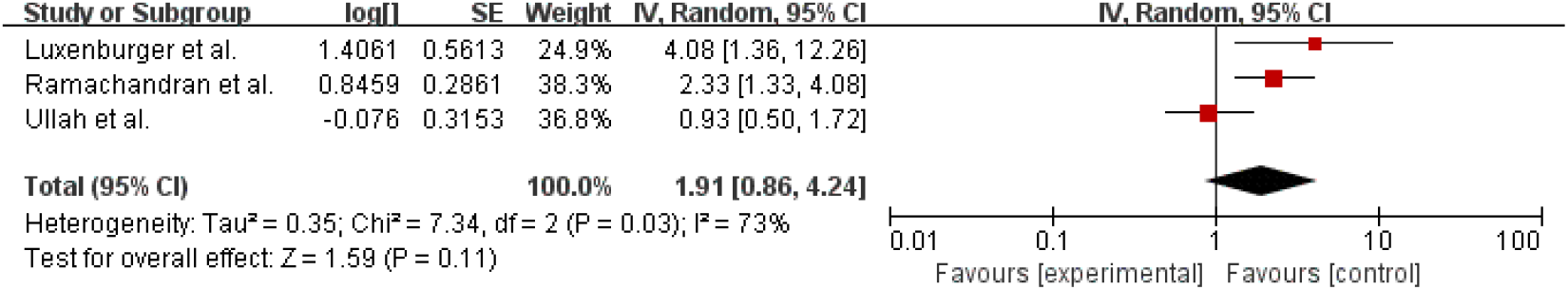
Forest plot: Association between PPI use and mortality risk for COVID-19 patients

#### Association between PPI use and risk of secondary infection for COVID-19 patients

Two articles^[12,17]^ investigating PPI use and risk of secondary infection were found in the database, we combined them and found that PPI use was associated with increased risk of secondary infection. (OR=4.33, 95% CI=2.57-7.29, P<0.001; *I*^*2*^=0%, *P*_*heterogeneity*_ =0.570) (**Figure 5**). Since there were only two publications in this group, a test for publication bias and sensitive analysis were not performed.

**Figure 5.**
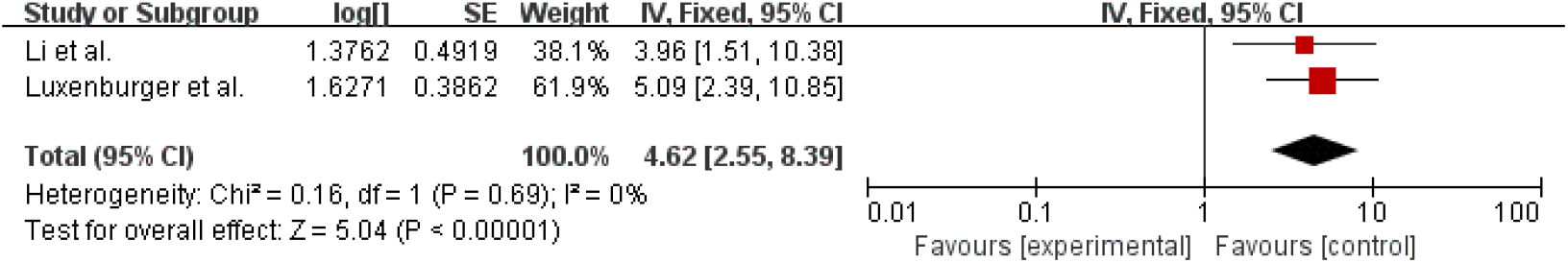
Forest plot: Association between PPI use and risk of secondary infection for COVID-19 patients

## Discussion

Overall, this meta-analysis included a total of fourteen original studies. Two original studies from a previous meta-analysis^[22]^ were excluded as they did not meet our inclusion criteria. In the study conducted by Yan et al^[23]^, the exposure for patients is acid-suppression drugs instead of PPI only, which may result in bias and obscure the result since acid-suppression drugs include not only PPIs proton pump but also other classes of medications, such as histamine receptor blockers. Moreover, Li et al^[22]^ categorized Almitrine from the study conducted by Losser et al^[24]^ as PPI, however, Almitrine is not PPI but a respiratory stimulant^[25]^. We performed analysis based on four different clinical outcomes. Firstly, PPI use is not linked to increased risk of COVID-19 infection, and subgroup analysis also found no increased risk in cohort studies and case-control studies, as well as Asian countries and western countries. This means patients in the general population who are taking PPIs may not need to stop taking it for fear of an increased risk of COVID-19 infection, same to the conclusion of a previous meta-analysis^[26]^. PPIs act through the inhibition of the hydrogen/potassium pump (H+/K+-ATPase pump) in the gastric parietal cells^[27]^. However, this strong inhibition of gastric acid secretion increases the alkalinity of gastric juice, thereby reducing the protective effect on the gastrointestinal tract, which might potentially help SARS-CoV-2 survive in the stomach and infiltrate GI epithelial cells^[4]^. Although risk of transmission through gastrointestinal tract could not be ruled out^[28]^, the most significant transmission route for SARS-CoV-2 is the respiratory route, including droplet transmission and aerosol transmission^[29-30]^, which may temper the effects of PPI on the gastrointestinal tract in terms of COVID-19 infection. Therefore, we need to be prudent when interpreting the current result of COVID-19 infection risk for PPI users.

Secondly, a 67% increased risk of serious illness in COVID-19 patients, including ICU admission, intubation, and death, was found for PPI users. Hence, the use of PPI should be carefully considered in patients infected with COVID-19, especially when prophylaxis for stress ulcer and gastrointestinal bleeding is needed for hospitalized patients. In subgroup analysis, the increased risk was found in both Asian countries and western countries, and was also observed in case-control studies, but in cohort studies, the increased risk was not statistically significant. Heterogeneity was found in the subgroup of PPI use and risk of severe disease, which is likely due to the criteria for defining severe illness varied among the included articles, such as admission to ICU, endotracheal intubation, and death. Despite the increased risk of severe illness, it is noteworthy that, there is no connection between PPI use and higher or lower risk of death in COVID-19 patients. This implies that it may not be necessary to discontinue PPI for patients simply for concern of increased mortality. However, in sensitivity analysis, the results of mortality changed significantly after exclusion of the study conducted by Ullah et al., suggesting it could be the source of the heterogeneity in this meta-analysis, as this study was conducted on patients with hepatobiliary and pancreatic disease instead of general population, and that the ORs were not adjusted based on multivariate analyses ^[11]^, therefore, the result of mortality risk of COVID-19 for PPI users should be interpreted cautiously.

Lastly, the subsequent risk of secondary infections is increased with the use of PPI. As mentioned above, PPI suppresses acid secretion^[27]^, and it has been shown that a decrease in gastric acid production may affect the gut microbial composition and increase the risk of intestinal infection^[27,31]^, including C*lostridioides difficile, Salmonella, Campylobacter* infection^[32,33]^. As previously reported, PPI use was also associated with greater risk of nosocomial pneumonia, especially in ICU patients, compared with histamine-2 receptor blockers^[34]^. Therefore, we need to be more cautious to use PPI for COVID-19 patients who already have high risk of secondary infection or who are immunocompromised. Nevertheless, only two original articles were included for assessing the risk of secondary infection, so the result needs to be interpreted with caution.

The inherent limitations of this study are as follows. First, due to insufficient information of original studies, more in-depth analysis on patient’s age, duration of PPI use and dose were not analyzed. Second, only two studies were included for the subgroup of secondary infection. Despite some of these limitations, the strengths of this study must be mentioned: First, no obvious publication bias in our study was detected. Second, compared with the previous meta-analysis, we included more literature and have more comprehensive subgroup analyses on different outcome index^[22,26,35]^.

In summary, although PPI use is not associated with increased risk infection and may not change the mortality risk of COVID-19, it appeared to be associated with increased risk of progression to severe disease, and increased risk of secondary infection. However, original studies included were limited, therefore more original studies to further clarify the relationship between PPI and COVID-19 are still urgently needed.

## Data Availability

This is a meta-analysis, and all data are extracted from original studies that can retrieved from online data bases and other resourses.

## Supportive foundations

No funding sources for this work.

## Author contributions

Dr. C. Yan developed search strategy, performed literature search, collected the data, performed statistical analysis, and wrote the manuscript. Y. Chen performed literature search, developed search strategy, collected the data, performed statistical analysis, and wrote the manuscript. Dr. C. Sun designed the study, developed search strategy, performed literature search, collected the data, performed statistical analysis and wrote the manuscript. Dr. M.A. Ahmed, Dr. C. Bhan and Dr. Chen provided critical opinion and revised the manuscript. Z. Guo, H. Yang, C. Cao, Z. Ji, Y Yan, and Y. Zuo participated in writing and revised the manuscript. H. Lei performed literature search and collected data. Dr Y. Sun and Dr. Y Li participated in literature search and data collection, and revised the manuscript. Dr. Q. Zhou provided critical opinion and revised the manuscript. All authors approved the final manuscript.

